# COVID 19 healthcare facility demand forecasts for rural residents

**DOI:** 10.1101/2020.06.05.20123380

**Authors:** Andrio Adwibowo

## Abstract

One of the main challenges in dealing with the current COVID 19 pandemic is how to fulfill the healthcare facility demands especially for the residents living in the rural areas that have restricted healthcare access. Correspondingly, this study aims to record the daily COVID 19 cases and continue with the forecasting of the average daily demand (ADD) of healthcare facilities including beds, ICUs, and ventilators using ARIMA model. The forecasts were made for 3 rural populations located in the southern Amazon. The model shows that the healthcare ADD was different in each population. Likewise, the model forecasts that in a rural population that has the highest daily case with projected average cases equal to 67 cases/day (95%CI: 24, 110), that population has to fulfill healthcare ADD consisting of 57 beds/day (95%CI: 21, 93), 8 ICUs/day (95%CI: 2, 14), and 2 ventilators/day (95%CI: 2, 3). To conclude, the ARIMA model has addressed critical questions about ADD for beds, ICUs, and ventilators for rural residents. This ARIMA model based healthcare plan will hopefully provide versatile tool to improve healthcare resource allocations.

## 1 Introduction

The COVID 19 pandemic has significantly affected all corners of the world, including rural areas (FAO 2020). These areas will be the next places vulnerable to COVID 19. Most of confirmed COVID 19 cases have been centered on major urban areas, nonetheless the cases now are shifting to smaller rural communities. While, majority of the people with low incomes and food insecurity live in rural areas and these conditions make them more vulnerable to severe pandemic impact. In fact in the developing world, rural residents are much less prepared to deal with the direct and impact of the pandemic.

Ranscombe (2020) has pointed out the COVID 19 pandemic risks on rural areas. First, the rural populations have much higher proportions of older people. Second, there are ill equipped and insufficient public healthcare facilities and fewer district hospitals. The other determinant factors observed in rural populations also including lack of awareness, a limited supply of clean water, and low levels of nutrition (Fontes *et al*. 1998).

Kumar *et al*. (2020) have provided comprehensive information regarding the current rural health facilities during COVID 19 pandemic. In their study they found rural health facility shortages. The percentages of the facilities at sub center, primary, and community levels were 18%, 22%, and 30% respectively. While those facility numbers have increased recently, nonetheless it is substantially below the World Health Organization standard. At the community levels, there were also a shortage of health specialists working including surgeons (84.6%), obstetricians and gynaecologists (74.7%), physicians (85.7%) and paediatricians (82.6%).

The rural health facility and specialist shortages were also observed in Africa (Maphumulo and Bhengu 2019) and South America (Massuda *et al*. 2018) continents. Naicker *et al*. (2009) reported that the medical schools were only available in 24 of the 47 countries in sub-Saharan Africa. Unlike in America that has 25 healthcare workers per 1000 population, in Africa only 2 healthcare workers available to support 1000 people.

One of the most important healthcares during the pandemic is the availability of hospital beds and intensive care units (ICU) which is expected to increase with the rising number of COVID 19 cases. It has been estimated that 65% of hospital and ICU beds are routinely occupied (Halpern *et al*. 2016). These equal to 277346 typically hospital beds and 34222 ICU beds. Correspondingly, rapidly rising COVID 19 cases in a country could overwhelm these limited healthcare resources and may cause impact mainly on the rural residents since they have limited access to those resources.

To deal with this challenge it is required a comprehensive planning of healthcare facility requirements including hospital and ICU beds. One of approaches that visible is by making estimations. Jombart *et al*. (2020) and Moghada *et al*. (2020) stated that developing an estimation of potential COVID 19 driven demands for hospital and ICU beds are fundamental and urgently required to plan operations dedicated to scaling up healthcare capacity during pandemic.

According to the rising COVID 19 cases and followed by potential demands of healthcare facilities as explained before, this study aims to forecast the healthcare requirement to deal with the COVID 19 cases mainly in rural settings. Since the healthcare demands and COVID 19 cases are presented as a function of time (day), an ARIMA model is used to develop the healthcare demand forecasting.

## 2. Methodology

### 2.1. Study populations

The study populations (Figure 1) were located in the southern Amazon and surrounded by the tropical rainforests and fragment of savanna and grassland (Sano *et al*. 2009, Galford *et al*. 2010). The populations included Cuiabá, Rondonópolis, and Várzea Grande that occupy the most of the savanna. The populations for Cuiabá, Rondonópolis, and Várzea Grande were 612547, 144049, and 184069 respectively.

**Figure 1.**
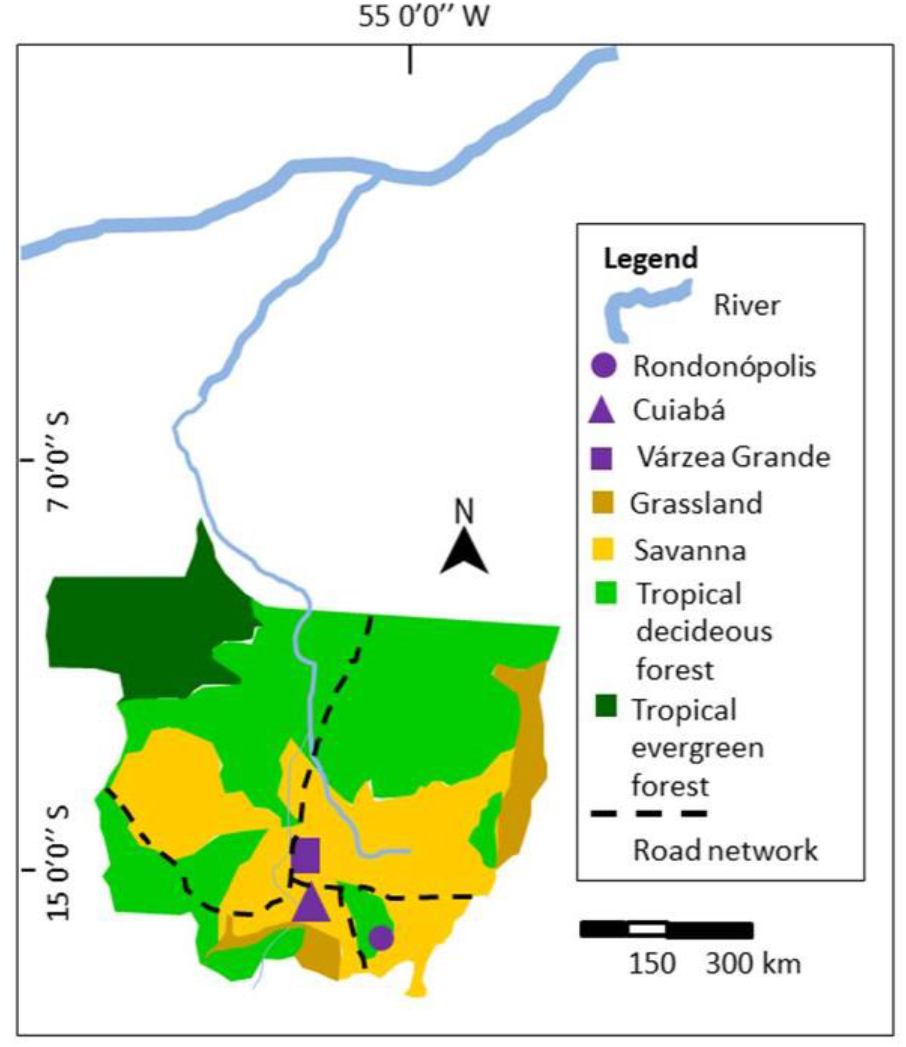
The study populations were located in the southern Amazon river.

### 2.2. Daily COVID 19 cases

Applied data used in this study consisted of daily and confirmed COVID 19 cases from study populations. The data was based on data for May 2020 obtained from Life Centre Institute, Indigenous Peoples Federation, and Council of Municipal Secretaries of Saúde de Matogrosso.

### 2.3. ARIMA forecasting

ARIMA is an abbreviation for Autoregressive Integrated Moving Average. This statistical approach is a versatile tool that had been used widely to forecast healthcare needs including disease management (Sato 2013), health expenditures (Ramezanian *et al*. 2019), and healthcare waste quantities (Chauhan and Singh 2017). In ARIMA, the autoregressive model is a statistical model expresses linear dependence between several time series. ARIMA estimates future of time series using its past and other series at several time lags.

ARIMA model has the statistical properties that make it best fitted for the forecasting of linear data patterns in this cases daily healthcare requirements developed from daily COVID 19 cases. It is based on the assumption of ARIMA model that the healthcare requirements of a variable are linear function of numerous past healthcare requirements and random errors. Likewise the fundamental process of generating a time series can be given in the following equation:

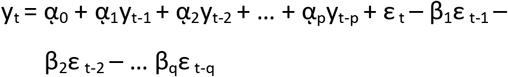

where γ_t_ and ε_t_ represents the actual value and random error at time period t, with α_i_(i = 1,2,3,…p) and β_j_(j = 1,2,3,…q) are model parameters. While p and q are integers that are usually referred as the order of the model. In addition, the ε_t_ which are random errors, are assumed to be independently and identically distributed with a mean of zero and a constant variance of σ^2^. The model then converges into an autoregressive (AR) model of order p, if q = 0 in above equation. Likewise the model converges into a moving average (MA) model of order q, if p = 0 in that equation. To conclude ARIMA model is functioning to decide an appropriate model of order p and q.

### 2.4. Healthcare facility demand estimation

The estimated healthcare facilities included the numbers of beds, ICUs, and ventilators required. The required healthcare facility number calculations were following Tyagi *et al*. (2020). Those required facilities were including isolation beds, ICUs, and ventilators for COVID-19 patients in the coming days. According to previous COVID 19 cases in other countries, the requirement of healthcare facilities increases as the infection reaches its peak. Likewise, the requirement estimations were developed based on predicted daily cases. The estimations were developed following the previous cases in China and Italy that healthcare facility requirements for isolation beds, ICUs, and ventilators supports were 85%, 10%, 5% of daily cases respectively. The calculations of health facilities as follows:

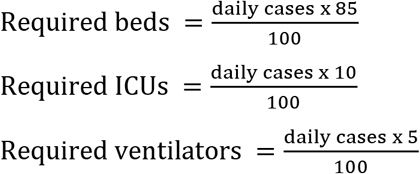

## 3. Results

Figure 2, 3, and 4 conveys about the actual confirmed and daily cases from 1^st^ to 30^th^ May 2020. The data shows that in all populations the cases have increasing trends. Currently, Cuiabá population has the highest case both for confirmed and daily cases.

**Figure 2.**
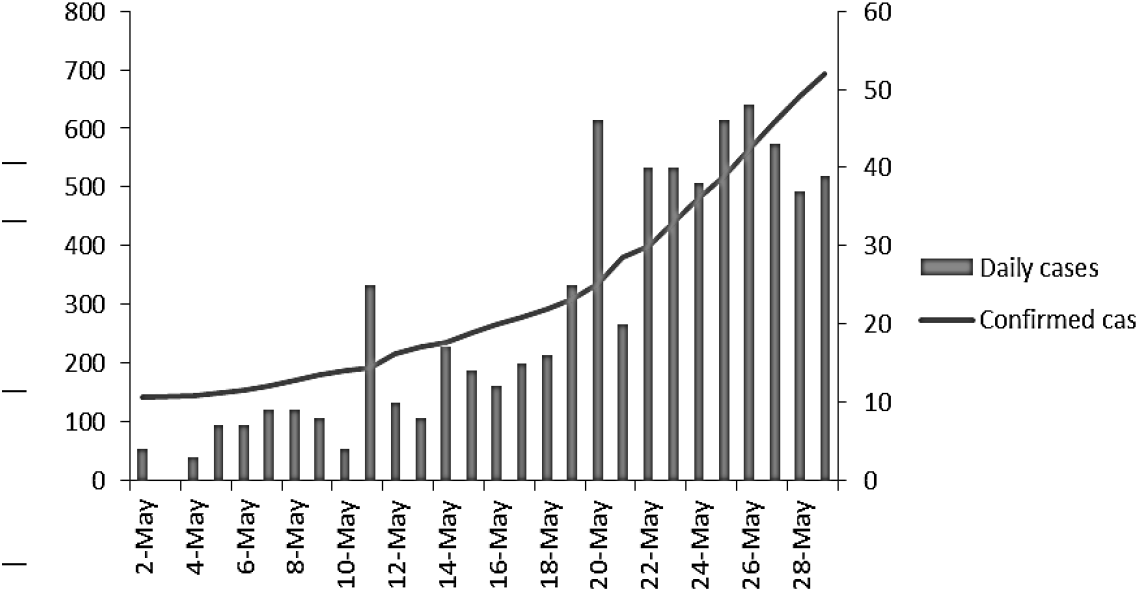
The confirmed and daily COVID 19 cases in Cuiabá population (confirmed cases on left y axis: 0-800, daily cases on right y axis: 0-60).

**Figure 3.**
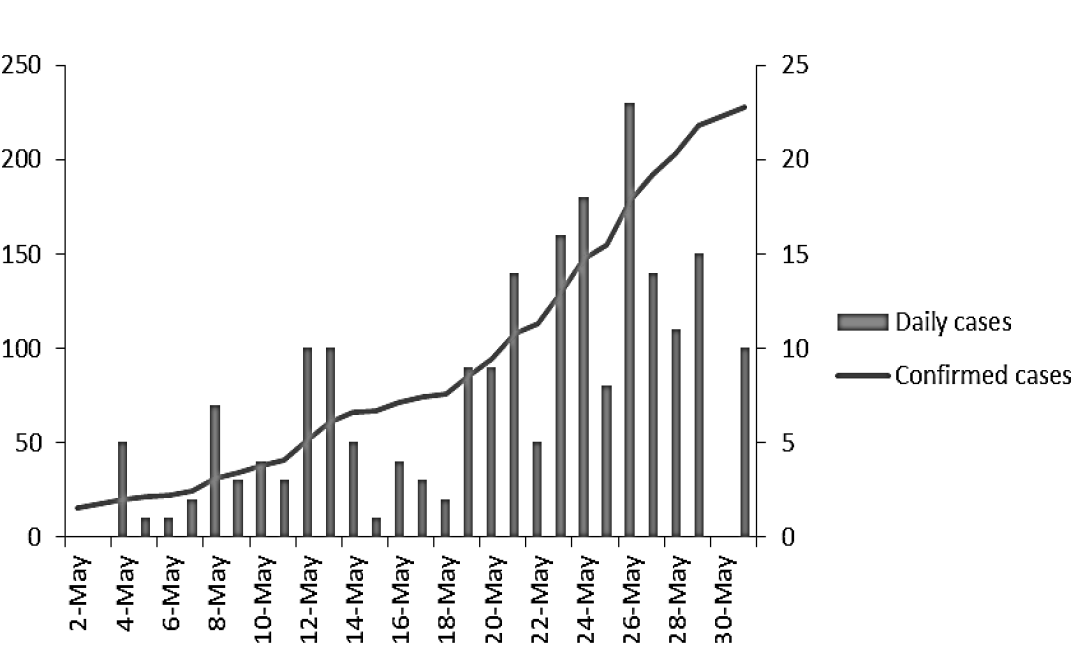
The confirmed and daily COVID 19 cases in Várzea Grande population. (confirmed cases on left y axis: 0–250, daily cases on right y axis: 0-25).

**Figure 4.**
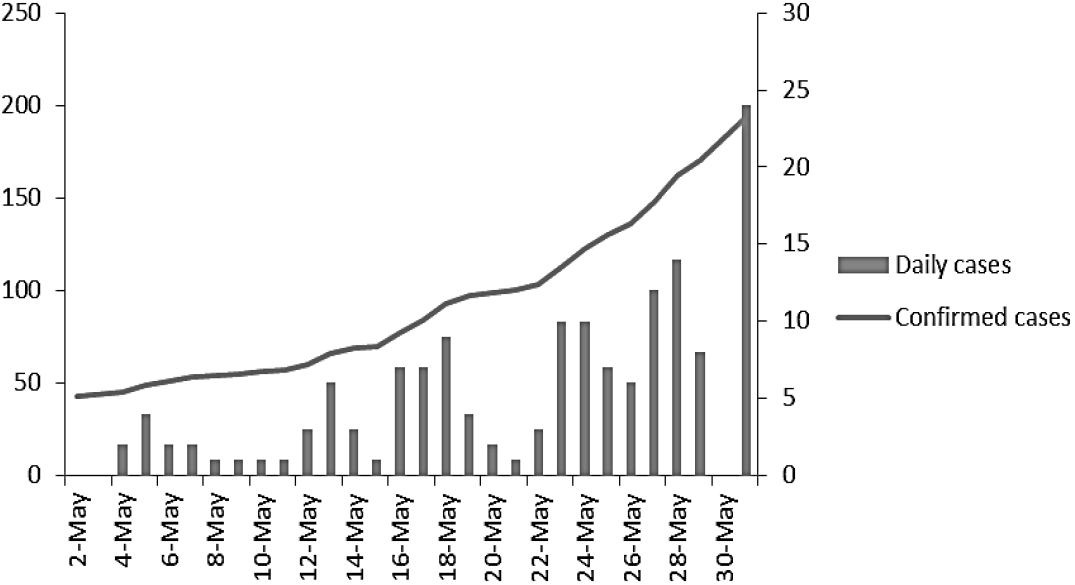
The confirmed and daily COVID 19 cases in Rondonópolis population. (confirmed cases on left y axis: 0–250, daily cases on right y axis: 0–30).

By aggregating estimations across populations, the overall trajectory of ARIMA model with a 95% confidence interval (shaded area) of expected daily healthcare facility requirements is shown in Figure 5, 6, and 7. The demand for healthcare facilities has increased rapidly after 30^th^ May. Requirement trajectory reflects the daily cases within each population. In Cuiabá population, the demand was estimated will increase rapidly. While the trends for healthcare facility demands in Várzea Grande and Rondonópolis were observed more stable and have slow growth as well.

**Figure 5.**
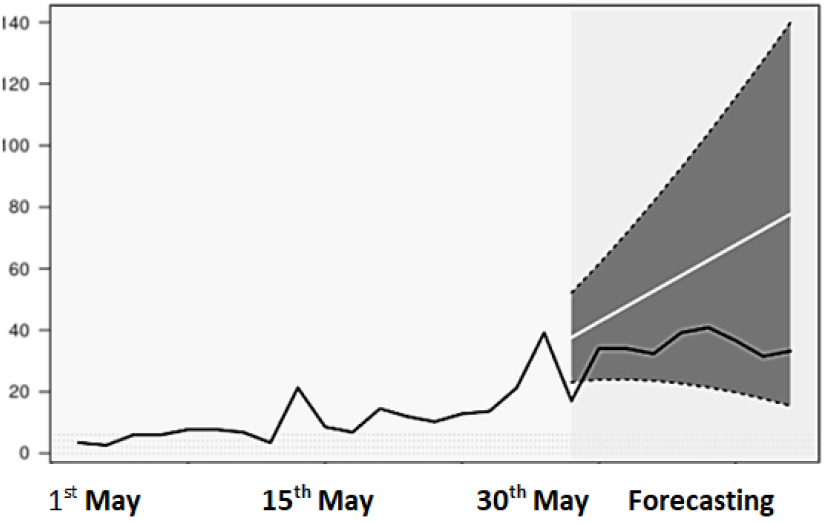
The estimated demands for beds/day with 95%CI (shaded area) in Cuiabá population.

**Figure 6.**
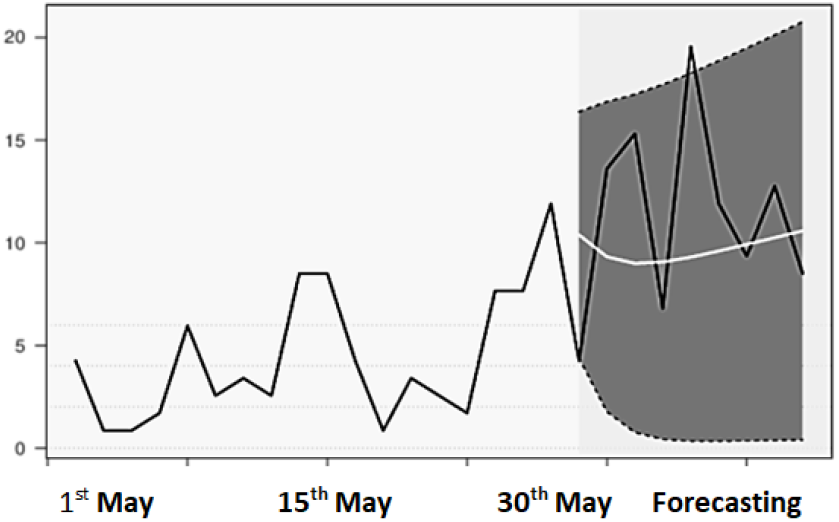
The estimated demands for beds/day with 95%CI (shaded area) in Várzea Grande population.

**Figure 7.**
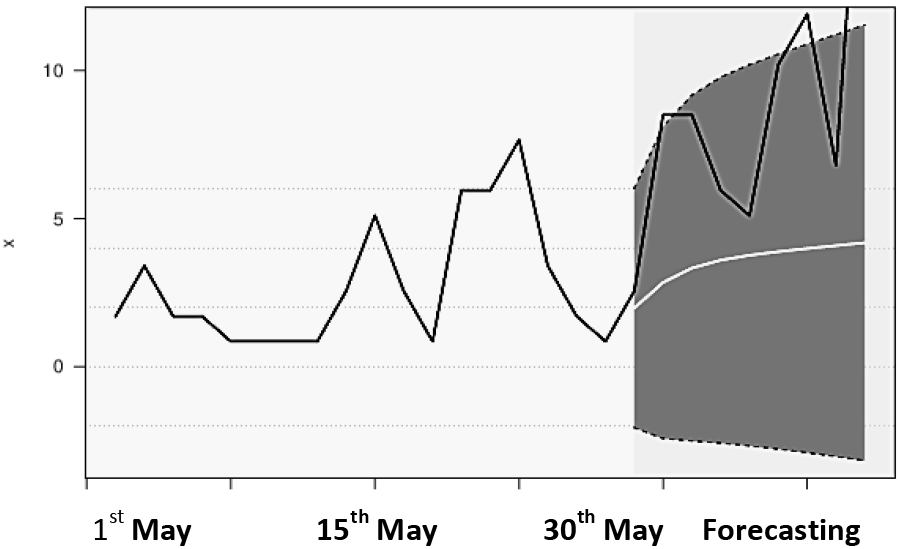
The estimated demands for beds/day with 95%CI (shaded area) in Rondonópolis population.

In a week after 30^th^ May in Cuiabá, the average daily demand (ADD) for healthcare facilities (beds, ICUs, ventilators) in Cuiabá is predicted to be 57 beds/day (95%CI: 21, 93), 8 ICU beds/day (95%CI: 2, 14), and 2 ventilators/day (95%CI: 2, 3). Based on the model, Cuiabá has the highest ADD of healthcare facility followed by Várzea Grande and Rondonópolis (Table 1, 2, 3). The lowest ADD was observed in Rondonópolis. In this population, it is estimated that the healthcare providers only require 3 beds/day (95%CI: −3, 10) and fewer ICU beds/day and ventilators/day compared to Cuiabá and Várzea Grande.

**Table 1.** Estimated average daily demand (ADD) for beds/day with 95%CI.

| Population | Mean | 95%low | 95%up |
| --- | --- | --- | --- |
| Cuiabá | 57 | 21 | 93 |
| Várzea Grande | 10 | 1 | 18 |
| Rondonópolis | 3 | <3 | 10 |

**Table 2.** Estimated average daily demand (ADD) for ICUs/day with 95%CI.

| Population | Mean | 95%low | 95%up |
| --- | --- | --- | --- |
| Cuiabá | 8 | 2 | 14 |
| Várzea Grande | 2 | 1 | 3 |
| Rondonópolis | <1 | <1 | 1 |

**Table 3.** Estimated average daily demand (ADD) for ventilators/day with 95%CI.

| Population | Mean | 95%low | 95%up |
| --- | --- | --- | --- |
| Cuiabá | 2 | 2 | 3 |
| Várzea Grande | 1 | <1 | 1 |
| Rondonópolis | <1 | <1 | <1 |

## 4. Discussions

Recently, there are large numbers of literatures have discussed the healthcare requirement projection using model during current COVID 19 pandemic (Rocklov 2020). The strength of this model lies on the use of parameters which can be measured. Even though in the onset of the virus outbreak, the parameter values cannot be identified and inserted. Nonetheless in the end, the parameters values can be identified from the actual data to reduce errors in model predictions. Since the model is developed using measurable parameters, the model calculations can be updated as the actual data of the parameters become available in the long run (Basu 2020).

Projected model is a versatile tool that can measure a healthcare provider capacity to fulfill the healthcare facility demand. Back to the day during SARS pandemics, ARIMA model was used to predict the number of occupied beds. The use of ARIMA model has enabled healthcare providers to predict 3 days ahead of time the number of required beds during the epidemic (Sato 2013).

A model can also identify whether in the future the healthcare facilities will be overwhelmed or not by the rising of COVID 19 cases. Zhang *et al*. (2020) have confirmed that based on the model, it was estimated that the COVID 19 cases have exceeded the available ICUs and ventilators within 21–27 days.

Estimated healthcare facility demand exceeding current capacity was also reported by Murray (2020). It has been estimated that in states that during pandemic peak, the demand for beds was estimated equal to 64175 (95%UI: 7977-251059) and this has exceeded 7% of all hospital beds available nationally. For ICU beds, the demand was 17380 (95%UI: 2432-57955). While the demand for ventilators was 19481 (95%UI: 9767-39674). The high upper bound of The 95% uncertainty intervals (UI) inform massive excess of the system.

Nonetheless, Tyagi *et al*. (2020) have warned that actual demand numbers can go higher than estimated healthcare infrastructures. The reasons were related to the people migrations. Unfortunately, people mobility could not be accounted for this projection model.

The model in this study shows that the healthcare facility requirements were different in each population. These differences are related to the numbers of confirmed cases in each population as well. This can be seen in Cuiabá population which has the highest case compared to other populations. The highest case in this population is followed by the highest projected healthcare facility as well. While the Várzea Grande and Rondonópolis populations that have lower cases are having lower requirements than Cuiabá. Ferstad *et al*. (2020) stated that a population with a high number of confirmed cases is likely to be first to see the demand for beds that even can exceed hospital beds availability.

The healthcare demands that can exceed the hospital beds availability in particular rural of southern Amazon have been concerned by Miranda *et al*. (2017). The concern was raised since there was a minimal available public hospital beds in the 12 rural Amazon cities to accommodate a surge in healthcare demands.

In contrast to the most current COVID 19 studies that have emphasized on the built urban inhabitants like large urban cities (Basu *et al*. 2020, Ha *et al*. 2020, Zou *et al*. 2020), this study is focusing on the rural inhabitants in particular savanna living ecosystems. This study is comparable to the other studies emphasizing on the healthcare conditions of rural inhabitants in savanna. The savanna due to its remoteness and geographic accessibility (Ihantamalala *et al*. 2020, Miguéis *et al*. 2018) may restrict access of inhabitants to the appropriate healthcare facilities. Cisse (2011) reported that numbers of health centers at the close proximity were low in the rural savanna and this has hindered residents to get vaccination and medical treatment at a lower cost. The rural resident activities which most of the residents’ livelihoods were depending on the agricultural sectors would pose a threat as well. During the start of the agricultural season, all residents will begin their farming activities in the pastoral fields. This may reduce the potential of disease transmitted among residents since the residents are spreading out from rural settlements. Nonetheless, in the pastoral field, the rural residents were in greater proximity without social distancing (Hughes and Hughes 2020) due to the collective works and this will increase their risks for infections (URD 2020).

## 5. Conclusions

This study has developed an ARIMA based model aims to facilitate healthcare planning in rural settings with estimation of the number of beds, ICUs, and ventilators necessary to support patients who require hospitalization for COVID 19 and how these compare to the available resources. The model estimates a daily time series demand for beds, ICUs, and ventilator requirements. To conclude, the ARIMA model has addressed critical questions about requirement estimations for beds, ICUs and equipment capacity such as ventilators. This planning model will hopefully provide an up to date tool for improved healthcare resource allocations and enable healthcare providers to create better management and short term predictions of the disease.

## 6. Recommendations

In response to the COVID 19 epidemic, healthcare providers especially in rural areas are urged to understand their current and future capacity to care the patients. Local rural leaders in a vulnerable rural area should pay particular attention to the case growth rate if they seek to tailor their response rapidly to debottleneck the healthcare facility constraints. Nonetheless the healthcare demands estimated in this study could be substantially higher if excess demand for healthcare resources is not addressed immediately. Regarding the model itself and for generating a more precise projection, in future the model environment can be easily updated with new parameter data.

## Data Availability

The data are available in the manuscript

## References

Basu S. 2020. Modelling to Predict Hospital Bed Requirements for Covid-19 Patients in California. medRXiv. 2020.05.17.20104919.

Chauhan A, Singh A. 2017. An ARIMA model for the forecasting of healthcare waste generation in the Garhwal region of Uttarakhand, India. International Journal of Services Operations and Informatics. 8(4).

Cisse A. 2011. Analysis of Health Care Utilization in Côte d’Ivoire. AERC Research Paper 201. FAO. 2020. COVID-19 and rural poverty:Supporting and protecting the rural poor in times of pandemic. Rome.

Ferstad JO, Gu A, Lee RY, Thapa I, Shin AY, Salomon JA, Glynn P, Shah NH, Milstein A, Schulman K, Scheinker D. 2020. A model to forecast regional demand for COVID-19 related hospital beds. medRxiv. 2020.03.26.20044842doi.

Fontes C, Rocha G, Rocha A, Mendes E, Queiroz D. 1998. Prevalence of Helicobacter pylori Infection in a Rural Area of the State of Mato Grosso, Brazil. Memórias do Instituto Oswaldo Cruz. 93(2).

Galford G, Melillo J, Kicklighter D, Cronin T, Cerri CE, Mustard J, Cerri C, Defries R. 2010. Greenhouse gas emissions from alternative futures of deforestation and agricultural management in the southern Amazon. Proceedings of the National Academy of Sciences of the United States of America. 107: 19649–19654.

Ha T, Schensul S, Lewis J, Brown S. 2020. Early assessment of knowledge, attitudes, anxiety and behavioral adaptations of Connecticut residents to COVID-19. medRxiv. 2020.05.18.20082073.

Halpern NA, Goldman KS, Tan S, Pastores M. 2016. Trends in critical care beds and use among population groups and medicare and medicaid beneficiaries in the United States: 2000–2010. Crit. Care Med. 44; 1490–1499.

Hughes RP and Hughes DA. 2020. Impact of relaxing COVID 19 social distancing measures on rural North Wales: a simulation analysis medRxiv. 2020.05.15.20102764.

Ihantamalala FA, Herbreteau V, Revillion C, Randriamihaja M, Commins J, Andreambeloson T, Rafenoarivamalala FH, Randrianambinina A, Cordier LF, Bonds MH, Garchitorena A. 2020.Improving geographical accessibility modeling for operational use by local health actors. medRxiv. 2020.03.09.2003310

Jombart T, Nightingale ES, Jit M, de Waroux O, Knight G, Flasche S, Eggo RM, Kucharski AJ, Pearson CAB, Procter SR, Edmunds JW. 2020. Forecasting critical care bed requirements for COVID-19 patients in England.

Kumar A, Nayar KR, Koya SF. COVID-19: Challenges and its consequences for rural health care in India. Public Health in Practice.

Maphumulo WT, Bhengu B R. 2019. Challenges of quality improvement in the healthcare of South Africa post-apartheid: A critical review. Curationis. 42(1): 1–9.

Massuda A, Hone T, Leles FAG, de Castro MC, Atun R. 2018. The Brazilian health system at crossroads: progress, crisis and resilience. BMJ Glob. Health. 3(4).

Miguéis G, da Silva RH, Guarim-Neto G, Damasceno Júnior GA.2018 Medicine bottled (garrafada): Rescue of the popular knowledge. Journal of Medicinal Plants Research. 12(22): 325–335.

Miranda ES, Shoaf K, Silva RS, Freitas CF, Osorio-de-Castro CGS. 2017. Expected hazards and hospital beds in host cities of the 2014 FIFA World Cup in Brazil. Cadernos de Saúde Pública. 33(5).

Moghada SM, Shoukat A, Fitzpatrick MC, Wells CR, Sah P, Pandey A, Sachs JD, Wang Z, Meyers LA, Singer BH, Galvani AP. 2020. Projecting hospital utilization during the COVID-19 outbreaks in the United States. Proceedings of the National Academy of Sciences. 117(16): 9122–9126.

Murray CJL. 2020. Forecasting COVID-19 impact on hospital bed-days, ICU-days, ventilator days and deaths by US state in the next 4 months. medRxiv. 2020.03.27.20043752v1.

Naicker S, & Plange-Rhule J, Tutt R, Eastwood J. 2009. Shortage of Healthcare Workers in Developing Countries—Africa. Ethnicity & disease. 19.

Ramezanian M, Haghdoost AA, Mehrolhassani MH, Abolhallaje M, Dehnavieh R, Najafi B, Fazaeli AA. 2019. Forecasting health expenditures in Iran using the ARIMA model (2016–2020). Medical journal of the Islamic Republic of Iran 33: 25.

Ranscombe P. 2020. Rural areas at risk during COVID-19 pandemic. Newsdesk 20(5):545.

Rocklov J. 2020. COVID-19 healthcare demand and mortality in Sweden in response to non-pharmaceutical (NPIs) mitigation and suppression scenarios. medRxiv 2020.03.20.20039594

Sano E, Rosa R, Brito J, Ferreira L. 2009. Land cover mapping of the tropical savanna region in Brazil. Environmental monitoring and assessment. 166: 113–24.

Sato RC. 2013. Disease management with ARIMA model in time series. Einstein (Sao Paulo). 11(1): 128–131.

Tyagi R, Bramhankar M, Pandey M, Kishore M. 2020. COVID 19: Real-time Forecasts of Confirmed Cases, Active Cases, and Health Infrastructure Requirements for India and its Majorly Affected States using the ARIMA model. medRxiv. 2020.05.17.20104588doi.

URD. 2020. COVID-19 in different contexts: north and south, urban and rural.

Zhang T, McFarlane K, Vallon J, Yang L, Xie J, Blanchet J, Glynn P, Staudenmayer K, Schulman K, Scheinker D. 2020. A model to estimate bed demand for COVID-19 related hospitalization. medRxiv. 2020.03.24.20042762doi.

Zou J, Bretin A, Gewirtz A. 2020. Antibodies to SARS/CoV-2 in arbitrarily-selected Atlanta residents. medRxiv 2020.05.01.20087478.

